# Reduced dengue incidence following deployments of *Wolbachia*-infected *Aedes aegypti* in Yogyakarta, Indonesia: a quasi-experimental trial using controlled interrupted time series analysis

**DOI:** 10.1101/2020.03.15.20036566

**Authors:** Citra Indriani, Warsito Tantowijoyo, Edwige Rancès, Bekti Andari, Equatori Prabowo, Dedik Yusdi, Muhammad Ridwan Ansari, Dwi Satria Wardhana, Endah Supriyati, Indah Nurhayati, Inggrid Ernesia, Sigit Setyawan, Iva Fitriana, Eggi Arguni, Yudiria Amelia, Riris Andono Ahmad, Nicholas P. Jewell, Suzanne M. Dufault, Peter A. Ryan, Benjamin R. Green, Thomas F. McAdam, Scott L. O’Neill, Stephanie K. Tanamas, Cameron P. Simmons, Katherine L. Anders, Adi Utarini

**Affiliations:** World Mosquito Program Yogyakarta, Centre of Tropical Medicine, Faculty of Medicine, Public Health and Nursing, Universitas Gadjah Mada, Yogyakarta, Indonesia; Department of Epidemiology, Biostatistics and Population Health, Faculty of Medicine, Public Health and Nursing, Universitas Gadjah Mada, Yogyakarta, Indonesia; World Mosquito Program, Institute of Vector Borne Disease, Monash University, Melbourne, Australia; Department of Paediatrics, Faculty of Medicine, Public Health and Nursing, Universitas Gadjah Mada, Yogyakarta, Indonesia; Yogyakarta City Health Office, Yogyakarta, Indonesia; Division of Epidemiology and Biostatistics, School of Public Health, University of California, Berkeley, CA, USA; Centre for Statistical Methodology, London School of Hygiene and Tropical Medicine, London, UK; Oxford University Clinical Research Unit, Hospital for Tropical Diseases, Ho Chi Minh City, Vietnam; Department of Health Policy and Management, Faculty of Medicine, Public Health and Nursing, Universitas Gadjah Mada, Yogyakarta, Indonesia

## Abstract

**Background:** *Ae. aegypti* mosquitoes stably transfected with the intracellular bacterium *Wolbachia pipientis* (*w*Mel strain) have been deployed for the biocontrol of dengue and related arboviral diseases in multiple countries. Field releases in northern Australia have previously demonstrated near elimination of local dengue transmission from *Wolbachia*-treated communities, and pilot studies in Indonesia have demonstrated the feasibility and acceptability of the method. We conducted a quasi-experimental trial to evaluate the impact of scaled *Wolbachia* releases on dengue incidence in an endemic setting in Indonesia.

**Methods and findings:** In Yogyakarta City, Indonesia, following an extensive community engagement campaign, *w*Mel *Wolbachia*-carrying mosquitoes were released every two weeks for 13–15 release rounds over seven months in 2016–17, in a contiguous 5 km^2^ area (population 65,000). A 3 km^2^ area (population 34,000) on the opposite side of the city was selected *a priori* as an untreated control area, on the basis of comparable socio-demographic characteristics and historical dengue incidence. Passive surveillance data on notified hospitalised dengue patients was used to evaluate the epidemiological impact of *Wolbachia* deployments, using controlled interrupted time series analysis. Rapid and sustained introgression of *w*Mel *Wolbachia* into local *Ae. aegypti* populations was achieved. Thirty-four dengue cases were notified from the intervention area and 53 from the control area (incidence 26 vs 79 per 100,000 person-years) during the 24 months after *Wolbachia* was deployed. This corresponded in the regression model to a 73% reduction in dengue incidence (95% confidence interval 49%,86%) associated with the *Wolbachia* intervention. Exploratory analysis including an additional 6 months of post-intervention observations showed a small strengthening of this effect (30 vs 115 per 100,000 person-years; 76% reduction in incidence, 95%CI 60%,86%).

**Conclusions:** These findings demonstrate a significant reduction in dengue incidence following successful introgression of *Wolbachia* into local *Ae. aegypti* populations in an endemic setting in Indonesia. These results are consistent with previous field trials in northern Australia, and support the effectiveness of this novel approach for the control of dengue and other *Aedes*-borne diseases.

## Background

*Aedes aegypti* mosquitoes are the primary vectors of dengue, Zika and chikungunya viruses. Of these, dengue is the most common arboviral infection of humans. Indonesia has among the highest dengue case burden, with an estimated 10 million clinical cases and 3000 deaths each year [1]. The annual per capita incidence is estimated at 36 - 44 symptomatic cases per 1000 population [1, 2] with a resultant cost in excess of USD $2 billion per annum [3]. Yogyakarta is typical of many cities in Indonesia; dengue is endemic with a seasonal peak between November and May. The high force of infection in Yogyakarta is evidenced by the hospitalised dengue case burden and high seroprevalence of dengue virus (DENV) neutralising antibodies (68%) in children 1 - 10 years [4].

The World Mosquito Program (WMP) is an international research collaboration deploying *Wolbachia*-infected *Ae. aegypti* mosquitoes for the biocontrol of dengue and other *Ae. aegypti*-borne viral infections. *Wolbachia* is an intracellular endosymbiotic bacterium present naturally in many insect species, but not present in *Ae. aegypti* until methods enabling stable transinfection emerged [5]. *Wolbachia* is maintained in *Ae. aegypti* through maternal inheritance, and confers a reproductive advantage via cytoplasmic incompatibility which facilitates its introgression into mosquito populations following open field release. A second feature of *w*Mel *Wolbachia* in *Ae. aegypti* is that it confers resistance to infection with all four DENV serotypes [6-9] plus other medically-important arboviruses like chikungunya, Zika and Yellow Fever [10-14]. Mathematical modelling predicts that the *w*Mel strain of *Wolbachia* should eliminate DENV transmission in most endemic country epidemiological circumstances [7, 15, 16].

The first open releases of *w*Mel *Wolbachia*-carrying *Ae. aegypti* were undertaken in northern Australia, in small isolated communities in 2011 and then into contiguous urban areas from 2013, resulting in the near elimination of local dengue transmission in northern Australia [17, 18]. In 2014-15, WMP conducted small-scale field trials in four small peri-urban communities (0.18–0.61 km^2^; populations 1157–2681) in Sleman and Bantul districts in Yogyakarta Province, Indonesia [19]. These proof-of-concept trials demonstrated successful introgression and long-term persistence of *Wolbachia* in local *Ae. aegypti* populations.

On the basis of these promising results, *w*Mel *Wolbachia*-carrying mosquitoes were deployed in the city of Yogyakarta in 2016–2017 with the dual aims of optimising methods for deployment at scale and demonstrating a reduction in arboviral disease incidence. Two consecutive prospective studies were conducted to evaluate the public health impact of *Wolbachia* releases in non-overlapping areas of Yogyakarta city (Figure 1). The first, reported here, was a quasi-experimental study in which interrupted time series analysis [20] of routine dengue surveillance data was used to evaluate the epidemiological impact of *Wolbachia* deployments in one contiguous area on the urban fringe, in comparison to a pre-specified untreated control area. The second study was a cluster randomised controlled trial (Applying *Wolbachia* to Eliminate Dengue (AWED); Clinicaltrials.gov #NCT03055585) [21]. Participant enrolment in the AWED trial is ongoing until late 2020, and no results are reported here.

**Figure 1.**
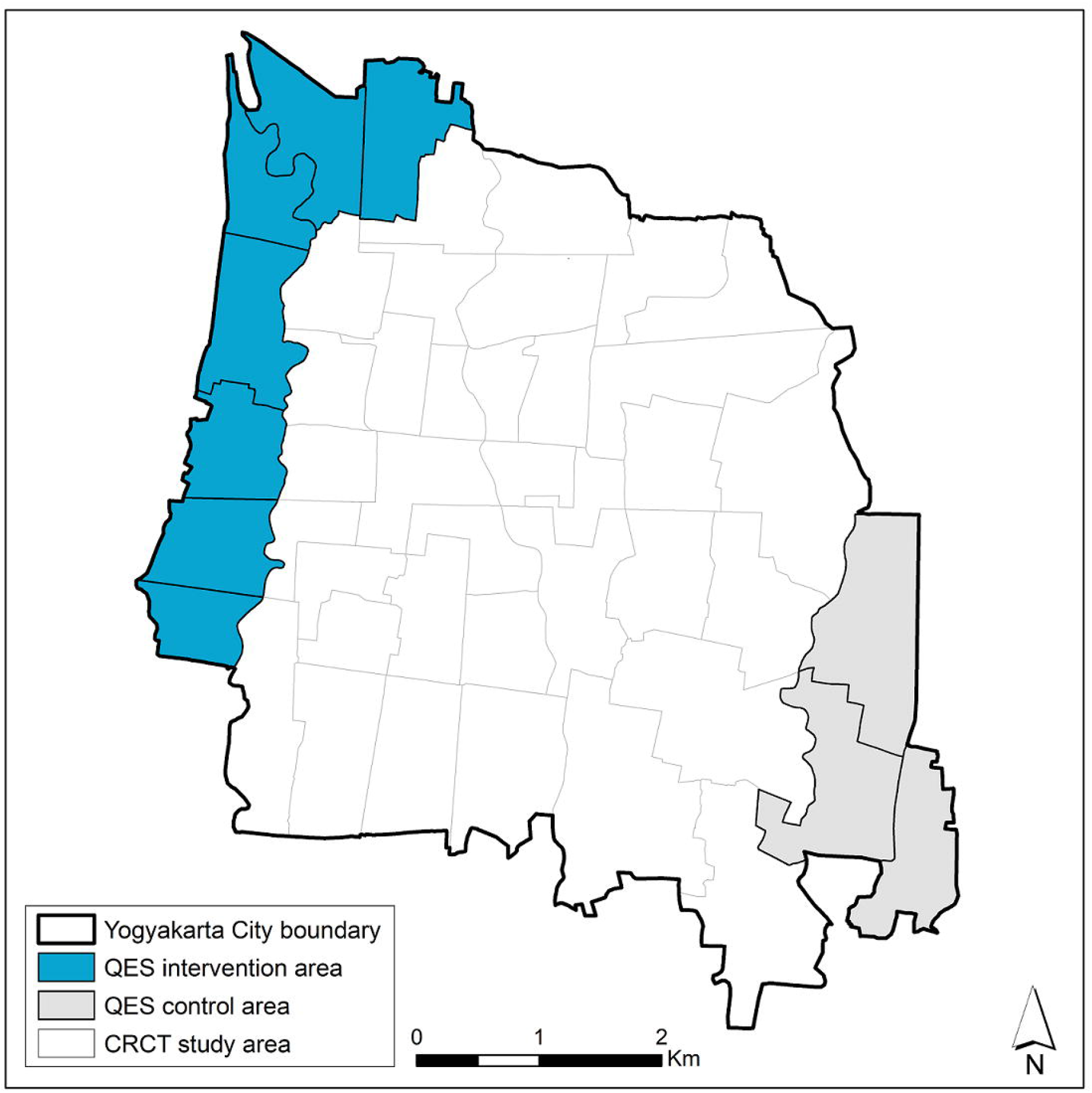
Map of intervention and control areas in the Yogyakarta quasi-experimental study (QES). The study area for the ‘Applying *Wolbachia* to Eliminate Dengue (AWED)’ cluster randomised controlled trial (CRCT), ongoing until late 2020, is also indicated.

The entomological and epidemiological results of the quasi-experimental study, at the *a priori* defined two year time point after *Wolbachia* deployment and with six months additional observation time, are reported here. They demonstrate a significant and sustained reduction in notified dengue haemorrhagic fever (DHF) case incidence in *Wolbachia*-treated communities.

## Methods

### Study setting

Yogyakarta City in south-central Java, Indonesia, has a population of 422,732 in an area of 32 km^2^, with 14 administrative districts comprising 45 kelurahans (urban villages). Seven kelurahans (total population 64,599; area 4.9 km^2^) on the north-western perimeter of the city were selected as the site for scaled deployment of *Wolbachia*-carrying mosquitoes (Figure 1), on the basis of logistical and operational feasibility. Three kelurahans (population 33,535; 3.1 km^2^) on the south-eastern perimeter were selected *a priori* as an untreated control area, on the basis of comparable socio-demographic characteristics (Table 1) and historical dengue incidence.

**Table 1:** Characteristics of intervention and control areas

| Area | Total size (km<sup>2</sup>) | Total population<sup>a</sup> | % completed high school<sup>b</sup> | % <15 years of age<sup>a</sup> |
| --- | --- | --- | --- | --- |
| Intervention (7 kelurahans) | 4.90 | 64 599 | 50% | 22% |
| Control (3 kelurahans) | 3.07 | 33 535 | 49% | 23% |

Source: <sup>a</sup>Statistics Indonesia (BPS) Yogyakarta City, 2017; <sup>b</sup>Yogyakarta Province Population Bureau, 2017.

### Community engagement

A Public Acceptance Model [17] was applied in engaging with the local community prior to *Wolbachia* deployments, and throughout the release and monitoring periods. Key elements of this approach included: meetings with key stakeholders and community leaders; meetings and ongoing regular communication with existing community reference groups at the village, city and provincial level; a communications campaign through social media, traditional media, mobile billboards, and community events; a household-based survey to evaluate awareness and acceptance prior to releases; and a ‘stakeholder enquiry system’ to receive and respond to any issues arising from stakeholders or community members.

A household survey of baseline community awareness of WMP’s *Wolbachia* method was undertaken in November 2015, followed by a second survey in June 2016 to evaluate awareness and acceptance prior to commencing releases. Two-stage cluster random sampling was used to select respondent households (n=587 in the 2015 awareness survey and n=862 in the 2016 awareness/acceptance survey). Administrative wards were the primary sampling unit, and 12 households were randomly selected per ward. The survey respondent was the head of household or another adult family member. Respondents were asked about their knowledge of the *Wolbachia* method, the primary source of information, their level of acceptance of *Wolbachia* releases in Yogyakarta City, their willingness to participate (host an MRC) if *Wolbachia* is implemented in their area, their support for *Wolbachia* as a method for controlling dengue in their community, their preferred method for receiving information about *Wolbachia* and their preferred method for consent. The survey participation rate was > 95% in each survey. During the 2015 baseline survey we found 15% (n=88) of households were aware of the *Wolbachia* technology, and this proportion increased in the 2016 pre-release survey to 22% (n=189). The most common source of information was community meetings. In the pre-release survey, 69% of households were accepting of the release of *Wolbachia*-infected *Ae. aegypti* as a new technology for controlling dengue, 6% were not accepting, and 25% neither agreed nor disagreed.

The stakeholder enquiry system was managed by a dedicated staff member and enquiries could be logged by phone call, SMS text, Whatsapp, email or direct communication with WMP field staff. In total we had 216 enquiries during the release phase, relating to: details of release activities (location, timing, methods) (35%), requests for face-to-face meetings with community leaders (25%), expressions of support for the *Wolbachia* technology (18%), reports of dengue cases in the release area (7%), reports from householders on the condition of mosquito release containers (MRCs) (6%), and refusal to host an MRC (6%).

### Mosquito rearing and egg production

An existing colony of local *Ae. aegypti* containing the *w*Mel *Wolbachia* strain, created in 2013 for pilot releases in Bantul District of Yogyakarta Province [19], was used as the founder colony for the releases described here. It was backcrossed for three generations with wild-type males collected from the study intervention area. Four cages with 150 wMel-carrying female *Ae aegypti* and 150 wild-type males were maintained for two gonotrophic cycles, and blood-fed by human volunteers. One hundred larvae were randomly selected from each cage and screened for *Wolbachia*, and only cages with 100% *Wolbachia* prevalence were maintained for the second backcrossing. An open colony was maintained by adding 10% of wild-type *Aegypti* after the second backcrossing. Insecticide resistance testing [22] demonstrated equivalent insecticide resistance profiles between the colony and wild material (Figure S1).

Mosquitoes were blood-fed by human volunteers once a week for two gonotrophic cycles, as per previous protocols [17]. All volunteer blood-feeders were afebrile and free of clinical signs or symptoms of any arbovirus infection both at the time of blood-feeding and for three days thereafter. Four days after blood feeding, mosquito eggs were collected by placing oviposition strips in each cage for 3 days. Eggs were dried slowly and stored in sealed containers until releases.

### Quality assurance of mosquito release material

For quality assurance (QA) of the mosquito colony, a sample of ∼1% of eggs from the parent generation of the release material was screened for *Wolbachia* by qualitative PCR Taqman assay on a Roche LightCycler 480. The pre-specified minimum acceptable *Wolbachia* prevalence was 97%. In each gonotrophic cycle, nine blood-fed female mosquitoes per human volunteer were tested for infection with DENV-1–4, chikungunya and Zika virus by qRT-PCR as previously described [23].

For QA of the release material, eggs from 10 oviposition strips were hatched each release week, with a minimum threshold hatch rate of 80%. Each release week, 10% of all release containers were selected randomly for QA, to estimate numbers of mosquitoes released. Eggs from these QA containers were manually counted under a microscope prior to field deployment. Release success was evaluated in the same QA containers at the time of servicing two weeks later (see below). A container was classified as ‘failed’ if: 1) it was lost; 2) it was dry; 3) pupae skin n<25;4) dead adult mosquitoes >10; 5) wing carcass >20; or 6) remaining larvae and pupae >30.

### Deployment

*Wolbachia*-carrying mosquitoes were released as eggs using mosquito release containers (MRCs). These were 2 litre plastic buckets each containing one oviposition strip with 100–150 eggs, Tetra Pleco Wafers fish food (Tetra GmbH, Germany), and 1 litre water. MRCs were covered and placed outside houses, protected from direct sun and rain. Holes drilled near the top of the bucket walls allowed adult mosquitoes to escape. Releases occurred between August 2016 and March 2017, with 13–15 rounds of releases in each kelurahan. Releases stopped in each kelurahan when the prevalence of *Wolbachia* in field-caught mosquitoes was >60% for three consecutive weeks releases (Table 2). MRCs were reset every two weeks. An MRC was placed in 1–2 randomly selected locations within each 50×50m grid square across the intervention area. Permission was obtained from property owners to place MRCs on private property.

**Table 2:** Summary of *Wolbachia* deployments in 7 urban villages

| Kelurahan<br>(urban village) | Total area<br>(km<sup>2</sup>) | Release start<br>and end date | # release<br>rounds | Mean release<br>points per round<br>(min, max) | Mean estimated<br>mosquitoes released<br>per round (st dev)<sup>a</sup> |
| --- | --- | --- | --- | --- | --- |
| Kricak | 0.84 | 15 Aug 2016 -<br>13 Feb 2017 | 15 | 315 (280, 362) | 23 756 (5337) |
| Pakuncen | 0.64 | 22 Aug 2016 -<br>6 Mar 2017 | 14 | 265 (193, 357) | 20 766 (9003) |
| Patangpuluhan | 0.45 | 24 Aug 2016 -<br>8 Mar 2017 | 14 | 189 (169, 233) | 14 401 (4440) |
| Tegalrejo | 0.82 | 7 Sept 2016 -<br>8 Mar 2017 | 13 | 260 (228, 312) | 18 786 (3852) |
| Bener | 0.59 | 5 Sept 2016 -<br>6 Mar 2017 | 13 | 126 (123, 133) | 9005 (1687) |
| Karangwaru | 0.77 | 15 Sept 2016 -<br>15 Mar 2017 | 13 | 269 (238, 314) | 21 564 (4426) |
| Wirobrajan | 0.79 | 18 Sept 2016 -<br>13 Mar 2017 | 13 | 260 (232, 320) | 19 691 (3949) |

<sup>a</sup> The number of mosquitoes released each round in each kelurahan was estimated from the mean number of eggs in the mosquito release containers (MRCs) randomly selected for quality assurance (QA), multiplied by the number of successful release containers in that kelurahan. Shown here is the mean number (and standard deviation) of mosquitoes released per release round, in each kelurahan.

### Monitoring

Prevalence of *Wolbachia* in the local *Ae. aegypti* population was monitored by weekly collection of adult mosquitoes via a network of 89 BG Sentinel traps (Biogents, Germany). The median (range) trap density was 15.7 (13.0–20.0) BG/km^2^ in the intervention area and 3.6 (2.0–4.0) BG/km^2^ in the non-release area. Mosquitoes were demobilised at -20°C for ≥1 hour, then identified by morphological features. The number of mosquitoes caught in each BG trap was recorded by species, sex, and in total. *Ae. aegypti* were stored at -20°C in 80% ethanol until *Wolbachia* screening.

### Diagnostics

Field-caught *Ae. aegypti* were screened for *w*Mel *Wolbachia* by qualitative PCR Taqman assay on a Roche LightCycler 480. Specific primers targeting the gene encoding *Ae aegypti Rps17* and *wMel WD0513* were used as previously described [24], but with replacement of the Cy5-BHQ3 fluorophore-quencher pair in the *w*Mel probe with the fluorophore-quencher LC640-IowaBlack (Integrated DNA technologies) [25]. Testing was weekly when *Wolbachia* prevalence was <80%; two-weekly when ≥80% and four-weekly when ≥90%.

### Epidemiological data

Data on hospitalized dengue haemorrhagic fever (DHF) cases (ICD-10 code A91; International Classification of Diseases 10th revision) were obtained from the Yogyakarta District Health Office (DHO), for January 2006 – September 2019. Data on hospitalised dengue fever (DF) cases (ICD-10 code A90) were also obtained for January 2017 – September 2019, as DF notification only began in 2017. Data were monthly case counts by kelurahan of residence. Population data were obtained from the Indonesian Bureau of Statistics. Anonymised results of dengue rapid diagnostic tests (RDTs) (SD Bioline Dengue Duo, Abbott, USA) performed in 18 primary health care clinics across Yogyakarta City from March 2016 – September 2019 were obtained directly from the clinics. RDT results were extracted for participants resident in the quasi-experimental study intervention or control area, based on the kelurahan of residence where available, otherwise the kelurahan of clinic location.

### Statistical analysis

The crude dengue incidence rate ratio in the pre-intervention and post-intervention periods was calculated as the aggregate number of DHF cases divided by the aggregate person-months, in the intervention area versus the control area. Crude dengue incidence was similarly calculated using an endpoint that included both DF and DHF case notifications, but only in the post-intervention period due to the lack of DF reporting prior to January 2017. The *Wolbachia* intervention effect was estimated using controlled interrupted time series analysis [17, 18, 20]. Negative binomial regression was used to model monthly DHF case counts in the aggregate intervention and control areas, with an offset for population size. The primary analysis, defined *a priori*, included data from January 2006 until March 2019, two years post-release. A secondary analysis included an additional six months of data to September 2019. Annual population estimates were used for 2006–2013 and 2015-2017; 2013 population estimates were used for 2014 and 2017 estimates were used for 2018 and 2019, due to unavailability of data for those years. Seasonal variability in dengue incidence was controlled using 6-monthly flexible cubic splines. A binary ‘group’ variable indicated the study arm (intervention or control). A binary ‘treatment’ variable distinguished the pre-intervention period (up to the end of *Wolbachia* deployments in the last release area) and the post-intervention period, and the intervention effect was estimated from the interaction between the ‘group’ and ‘treatment’ variables. This allows explicitly for a level change in the outcome (dengue case incidence) in both intervention and control arms in the post-intervention period, for example in a scenario where other secular effects coincident with the *Wolbachia* deployments may have influenced dengue incidence independently of *Wolbachia*.

The unavailability of DF data prior to January 2017 precluded an ITS analysis with a combined DF/DHF endpoint. Robust standard errors were used for all models. Analyses were performed using Stata® statistical software package version 14.2 (StataCorp, College Station, TX).

### Power calculation

Power was estimated using 1000 simulated datasets drawn from a negative binomial distribution fitted to an 11-year time series (2006–2016) of monthly DHF case notifications from the intervention and control areas prior to *Wolbachia* deployment. Post-intervention time periods of 1, 2, 3 or 4 years were simulated, with the pre-intervention period fixed at 7 years (maximum total simulated time series of 11 years). Dengue case numbers were not modified for the untreated control area, and for the *Wolbachia* intervention area were either kept at baseline values (for the simulation at the null; ie RR=1) or reduced proportionately (for simulations of intervention effects of RR=0.7, 0.6, 0.5, 0.4, 0.3). For each of these five ‘true’ effect sizes, applied to each of the 1000 simulated time series, the ‘observed’ effect size was calculated from a negative binomial regression model of monthly case counts in the intervention and control areas, as described above. Power was estimated from the proportion of 1000 simulated scenarios in which a significant intervention effect (p<0.05) was observed (Figure 2). With seven years of pre-intervention data and one year post-intervention follow-up, there was 80% power to detect a ≥30% reduction in dengue incidence (IRR=0.7). There was only a marginal gain in power from >1 year of follow-up, if the true reduction in dengue incidence is greater than 30%. Power calculations were performed using R (R Foundation for Statistical Computing, Austria) with the “MASS” package [26] used for estimation of the regression models.

**Figure 2.**
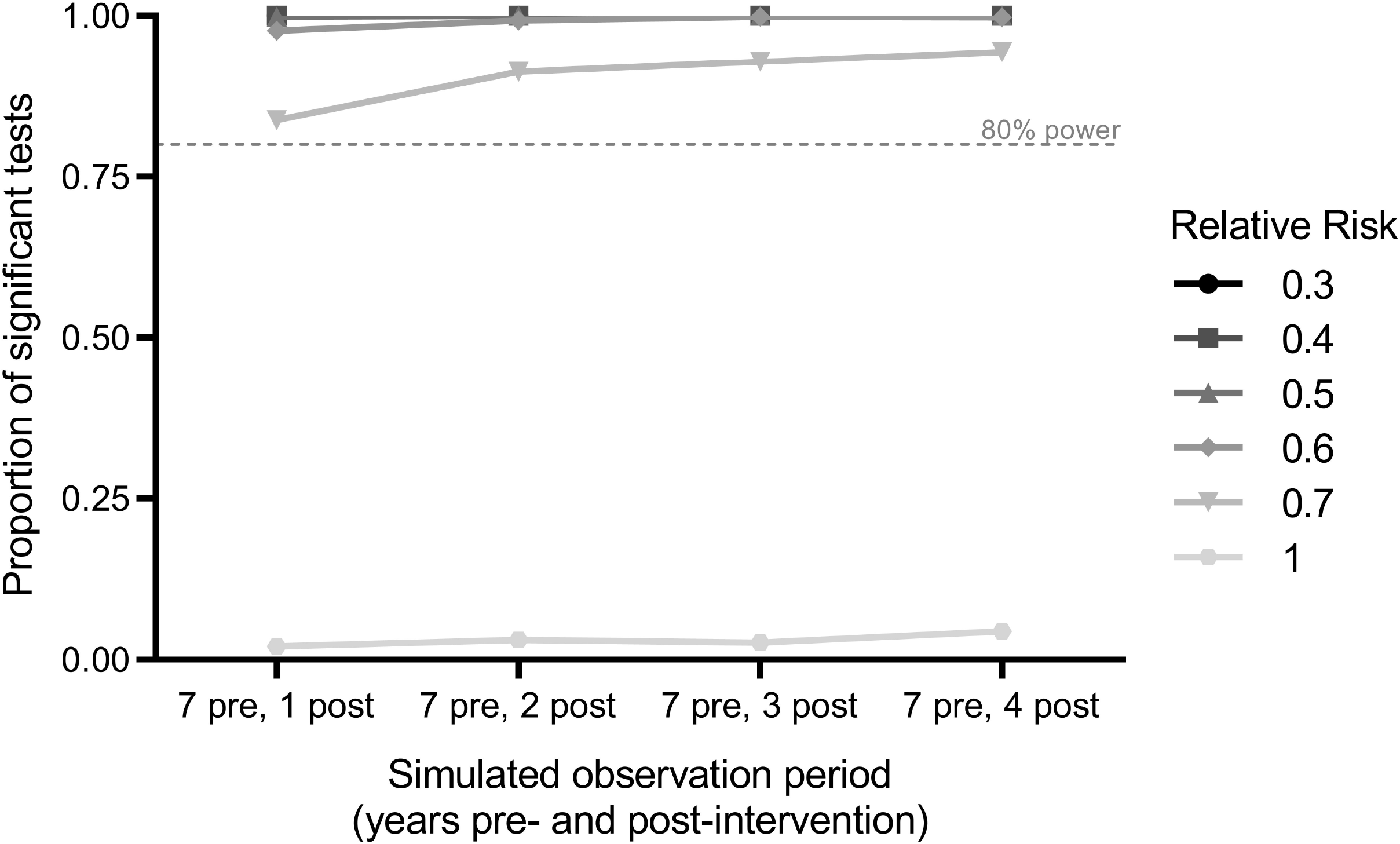
Power estimation. Power to detect a *Wolbachia*-associated reduction in dengue incidence using interrupted time series analysis was calculated as the proportion of significant results out of 1,000 simulations for varying post-intervention observation periods and relative risks.

### Regulatory and ethics approval and consent

An independent risk assessment conducted by the Indonesia Ministry of Research, Technology and Higher Education in 2016 concluded that over the next 30 years, there is a negligible risk of harm as a result of releasing *Wolbachia*-infected *Ae*.*aegypti* [27].

Approval to release *Wolbachia* mosquitoes was obtained from the provincial and city governments of Yogyakarta prior to releases. Ethical approval was obtained from the Universitas Gadjah Mada Faculty of Medicine, Public Health and Nursing Ethics Committee (approval number KE/FK/105/EC/2016) to release *Wolbachia* mosquitoes, for blood-feeding the mosquito colony on human volunteers, and to access non-identifiable aggregate data on monthly notified DHF and DF case numbers from the Yogyakarta DHO. Written agreement to share dengue surveillance data and non-identifiable individual dengue RDT results from puskesmas clinics was obtained from Yogyakarta DHO.

Verbal and written consent was obtained from heads of households for hosting a BG trap or MRC, and BG hosts were compensated for the cost of powering traps.

## Results

### *Wolbachia* establishment

Longitudinal entomological monitoring demonstrated rapid establishment of *Wolbachia* in the intervention areas, a continuous increase in *Wolbachia* prevalence in trapped *Ae. aegypti* throughout the first year post-release, and persistence at a very high prevalence ever since (Figure 3). The median *Wolbachia* prevalence was 73% (range 67–92%) one week after releases stopped, and 100% (96–100%) two years post-deployment. In the control areas, single *Wolbachia*-infected *Ae. aegypti* mosquitoes were detected on 11 occasions, but there has been no evidence of *Wolbachia* establishment.

**Figure 3.**
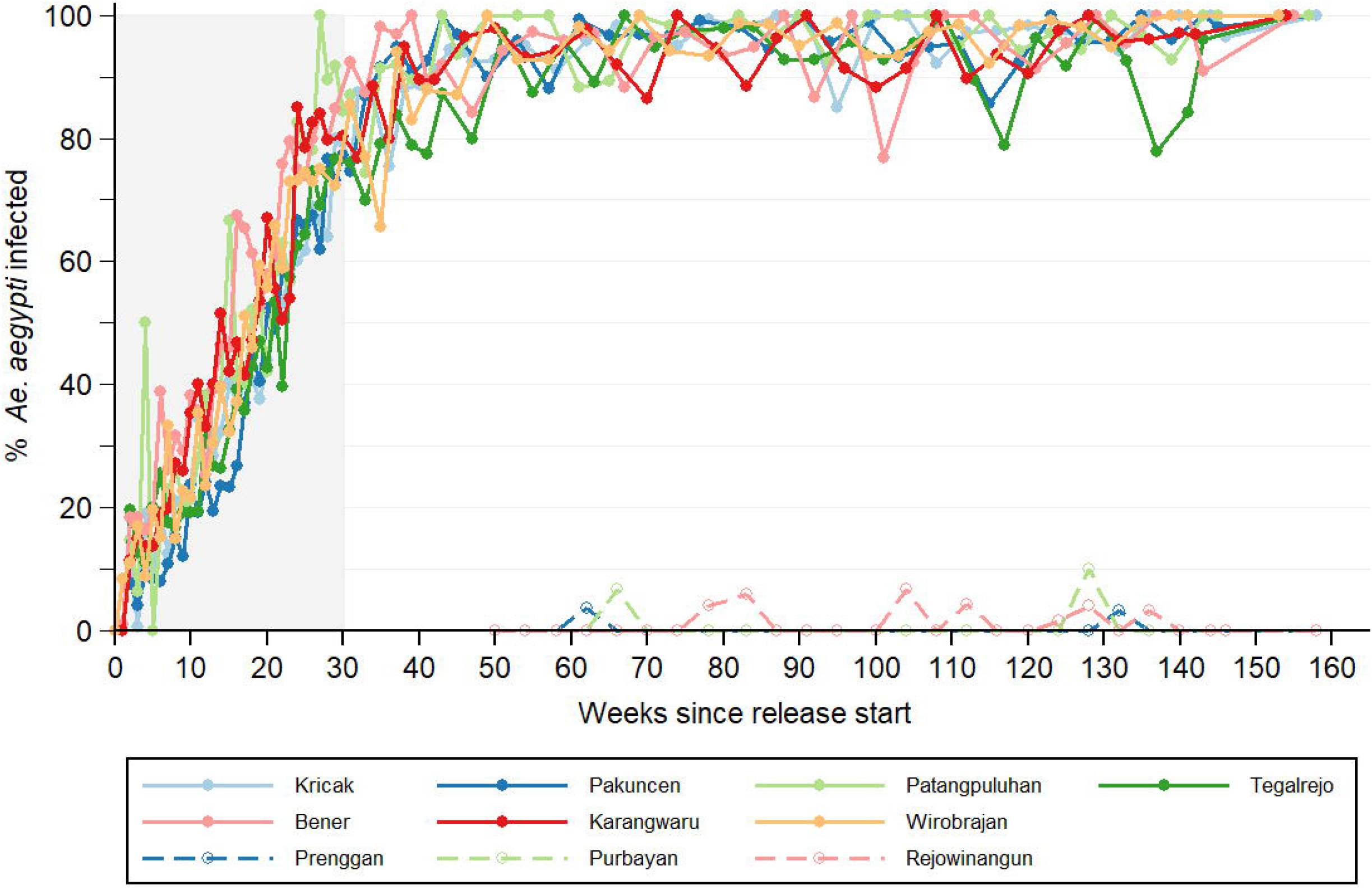
*Wolbachia* infection prevalence in local *Aedes aegypti* mosquito populations. Lines show the percentage of *Aedes aegypti* collected from intervention areas (closed circles; solid line) and untreated control areas (open circles; dashed line) that *Wolbachia* infected, each week since the start of deployments until September 2019. For the intervention areas, week 0 is the week in which deployment commenced (between 15 August and 18 September 2016, see Table 2). For the control areas, week 0 is the week in which the first deployments commenced in the intervention area (15 August 2016). Shaded area indicates release period.

### Dengue incidence in intervention and control areas

In the decade prior to *Wolbachia* releases, dengue outbreaks occurred annually in both the intervention and control areas (Figure 4). A median of 125 DHF cases were notified each year in the intervention area (range 65–308), corresponding to a median annual incidence of 169 per 100,000 population (range 98–477 per 100,000). In the control area, a median of 44 DHF cases were notified each year (range 19–159) corresponding to a median annual incidence of 130 per 100,000 (range 59–477 per 100,000). Per-capita dengue incidence in the intervention area was on average 15% higher than the control area during the pre-intervention period (crude IRR 1.15, p = 0.002). Monthly dengue incidence in the intervention and control areas was highly correlated over time (Spearman’s rho = 0.75, p<0.001), supporting the validity of the control area as a counterfactual for evaluating the epidemiological impact of *Wolbachia* releases (Figure 4).

**Figure 4:**
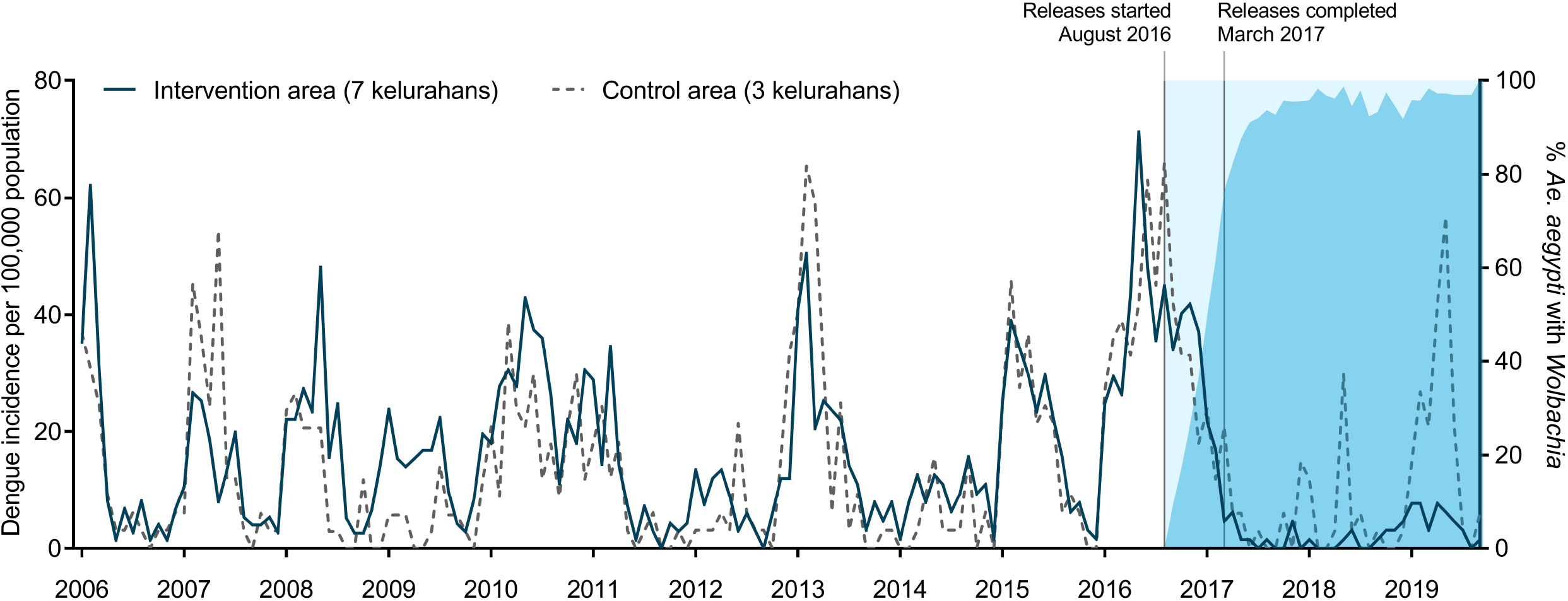
Dengue incidence in intervention and control areas, before and after the *Wolbachia* intervention. Monthly notified dengue case incidence (per 100,000 population) in the intervention (solid line) and control (dashed line) areas before and after *Wolbachia* deployments, January 2006 - September 2019. Blue shading indicates the *Wolbachia* infection prevalence in *Ae. aegypti* collected from the intervention area.

### *Wolbachia*-associated reduction in dengue incidence

During two years following the completion of *Wolbachia* deployments (April 2017 – March 2019), 34 DHF cases were notified from the intervention area and 53 from the control area, corresponding to 67% lower crude incidence in the *Wolbachia-*treated area (26 vs 79 cases per 100,000 person-years; IRR = 0.33, p<0.001).

In an interrupted time series (ITS) analysis of monthly DHF case notifications January 2006 – March 2019, which adjusts for baseline differences between the intervention and control areas and seasonal and inter-annual time effects, this translates to a 73% reduction in notified DHF incidence (95% CI 49%,86%; p<0.001) associated with the *Wolbachia* intervention.

In an exploratory analysis including six months of additional post-intervention data to September 2019, the DHF incidence in intervention vs control areas was 30 vs 115 per 100,000 person-years (crude IRR = 0.26, p<0.001) which translates in the regression model to a 76% reduction in dengue incidence (95% CI 60%,86%; p<0.001) associated with the *Wolbachia* intervention

During the first 2.5 years post-intervention (April 2017 – September 2019), 97 cases (DF + DHF) were notified from the intervention area and 137 from the control area (60 vs 163 per 100,000 person-years; IRR = 0.37, p<0.001) corresponding to 63% lower crude incidence in the *Wolbachia*-treated area, using a combined endpoint of DF and DHF case notifications. The lack of pre-release data on hospitalised DF cases precluded an ITS analysis with this combined endpoint.

### Dengue diagnostic findings in intervention and control areas

Prior to *Wolbachia* deployments (March 2016 – March 2017), there was no difference between intervention and control areas in the proportion of tested patients with an NS1-positive RDT result (127/447 (28%) in intervention area vs 106/408 (26%) in control area; Fishers exact p=0.44; Figure 5). By comparison, in the post-intervention period (April 2017 – September 2019) NS1 positivity was significantly lower in the intervention area than the control area (6/568 (1%) vs 61/429 (14%); Fishers exact p<0.001; Figure 5).

**Figure 5:**
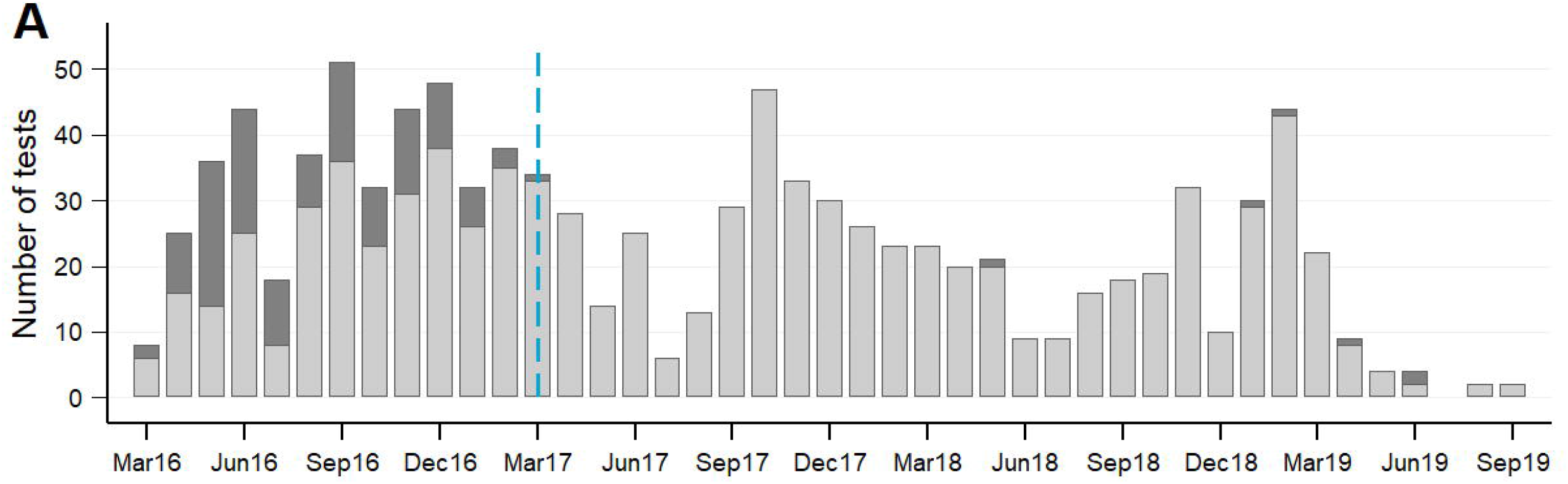
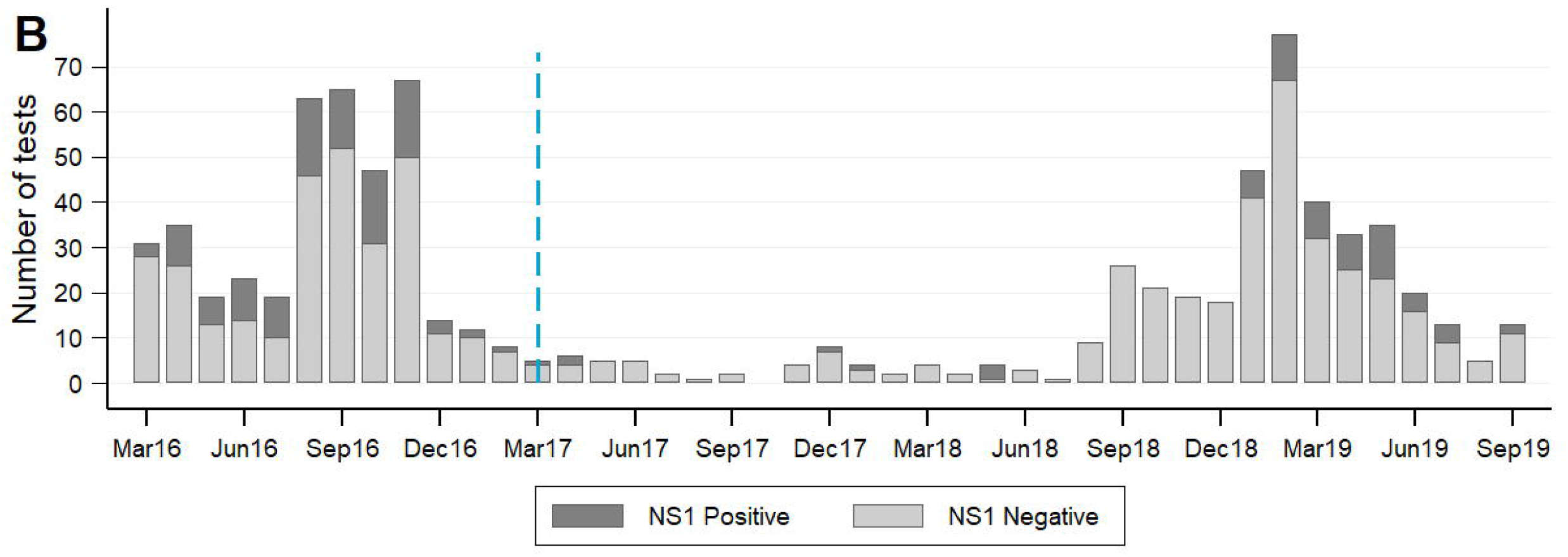
Laboratory-confirmed dengue cases in intervention and control areas, before and after the *Wolbachia* intervention. Dengue rapid diagnostic test results for patients presenting to primary care clinics in the intervention (A) and control (B) areas. Standard Diagnostics Dengue Duo rapid diagnostic kits for the detection of dengue virus NS1 antigen and IgM/IgG antibody were available in primary care clinics throughout Yogyakarta city from March 2016, and were used at the discretion of clinic staff as part of routine clinical care. Monthly counts of positive and negative results for DENV NS1 antigen, as recorded by clinic staff, were aggregated for all patients resident in the intervention area or control area. The blue dashed line indicates the completion of *Wolbachia* releases in the intervention area in March 2017.

## Discussion

Enabled by a successful community engagement campaign, release of *w*Mel *Wolbachia*-carrying mosquitoes throughout an urban community of 65,000 people in Yogyakarta City, Indonesia, resulted in rapid introgression and durable establishment of *Wolbachia* in the local *Ae. aegypti* population. Predefined analysis of public health surveillance data on dengue case notifications demonstrated a 73% reduction in DHF incidence in the intervention area during the 2 years after *Wolbachia* deployment, compared to a counterfactual untreated control area with a comparable historical incidence of disease. These epidemiological data from a dengue endemic setting are consistent with previous positive public health findings in Australia [17, 18].

Nazni *et al* recently reported on *w*AlbB establishment in *Ae. aegypti* in Kuala Lumpur, Malaysia including results of an exploratory analysis suggestive of a modest reduction (40%, 95%CI 5– 65%) in dengue incidence in release areas compared to several untreated control areas [28]. Nazni *et al* elected to use the *w*AlbB strain on the basis of a better thermostability profile than *w*Mel in the mosquito larval life stages. However we found no evidence that *w*Mel establishment was compromised in the climatic conditions of Yogyakarta. Elsewhere Ross *et al* reported that heatwaves have only transient effects on *w*Mel frequencies in Australia [29]. Evidence from pilot field studies is likely the best method for selecting the optimal *Wolbachia* strain for deployment in a given setting.

By chance, the commencement of *Wolbachia* releases in the quasi-experimental study area in late 2016 coincided with the largest dengue epidemic on record in Yogyakarta City.

Unsurprisingly, dengue case notifications to the District Health Office from across Yogyakarta City were then lower during 2017 and 2018 than in any two-year period in the previous 25 years, and it is notable that a highly significant *Wolbachia*-associated effect was observed even during this period of record low dengue transmission. A resurgence in dengue incidence was seen throughout southeast Asia in 2019 [30], including in Indonesia. Our exploratory analysis including an additional six months of observations after the *a priori* analysis time point of March 2019 showed a small strengthening of the intervention effect and tighter confidence intervals around the point estimate. Evidence of field effectiveness will continue to accumulate throughout subsequent dengue epidemic seasons. Our *a priori* analysis plan includes re-estimation of the intervention effect every 12 months, until five years post-intervention.

The dengue case time series used to evaluate the effectiveness of *w*Mel *Wolbachia* included only hospitalised patients with a clinical diagnosis of DHF, which was historically the case definition for mandatory notification in Indonesia. Using additional notifications data available since January 2017 for patients hospitalised with a clinical diagnosis of dengue fever (DF), we saw moderate attenuation of the intervention effect compared with the endpoint of DHF alone (63% vs 74% lower crude incidence in intervention area). Rather than a real interaction between the intervention effect and disease severity, we hypothesise that this reflects the reduced specificity of the DF clinical case definition compared with DHF, and the limited usage of confirmatory diagnostic tests; i.e. a greater proportion of notified DHF cases than DF cases represent true DENV infections. The hospitalised dengue patient population represents a more severe clinical subgroup of a much larger clinical burden, most of which is not counted in disease surveillance systems. Estimates of expansion factors from notified hospitalised cases to the true case burden in Indonesia range from 7–11 [31]. The AWED CRCT currently underway in Yogyakarta will address whether the incidence of ambulatory dengue cases, who are usually not detected by passive dengue case surveillance systems, is also reduced by *Wolbachia*.

The results reported here are consistent with mathematical modelling projections. Those predictions suggested that *w*Mel introgression would eliminate DENV transmission in most endemic settings for over a decade [7, 32]. A recent modelling study estimated that Indonesia had 7.8 million (95% uncertainty interval 1.8–17.7 million) symptomatic dengue cases in 2015, which were associated with 332,865 (94,175–754,203) lost disability-adjusted life years [16]. The authors estimated that a nationwide *w*Mel *Wolbachia* campaign could avert a large proportion of this burden (86.2%, 36.2–99.9%), with elimination predicted in low transmission settings. There are several reasons why elimination of DHF case notifications in the *Wolbachia* intervention area was not observed in the present study, and was likely infeasible. First, the geographic scale of the intervention area (∼5 km^2^) coupled with the mobility of the resident population means that some notified cases could have acquired their DENV infection at a location outside of the *Wolbachia*-treated area. Second, the specificity of DHF case reporting is likely to be imperfect, such that some DHF case notifications were not true dengue cases. Third, the intervention may not be entirely efficacious in blocking transmission of all DENV serotypes across the spectrum of transmission intensities that occurred during the period of observation.

Our study had limitations. The short-term, but open label nature of releasing *Wolbachia*-infected mosquitoes means it is plausible that community awareness altered health-care seeking behaviour, leading to differences in DHF notifications. However we believe this is unlikely to explain such a large and prolonged difference in hospitalised DHF incidence. We also cannot exclude a concurrent change in dengue control practices between the intervention and control areas, although we are not aware of any such alterations to standard practice. Due to the pragmatic nature of the study we did not determine the DENV serotypes circulating during the study period nor the incidence of other *Ae. aegypti-*borne diseases like Zika and chikungunya. The AWED CRCT currently underway in Yogyakarta City [21] should provide further detailed evidence on the impact of *w*Mel *Wolbachia* on the incidence of dengue, individual DENV serotypes and other arboviral diseases.

This study demonstrated the feasibility, acceptability and positive public health impact of the *w*Mel *Wolbachia* introgression method in a community of 65,000 people in Yogyakarta, Indonesia. This represents the first field evidence from an endemic setting of the effectiveness of this novel strategy in reducing dengue incidence, and is consistent with previous results from northern Australia where *Wolbachia* deployments have resulted in the effective elimination of local dengue transmission. Additional epidemiological evidence from this quasi-experimental study, and also the AWED trial, will accumulate during 2020/21. Continued optimisation of *Wolbachia* deployments plus additional long-term safety and efficacy data from a range of ecological and transmission settings will further enhance the attractiveness of this approach for disease control efforts.

## Data Availability

The entomological datasets used during the current study are available from the authors on reasonable request. The dengue case notification data and NS1 rapid diagnostic test results are available from the Yogyakarta District Health Office and individual Yogyakarta puskesmas clinics, respectively, but restrictions apply to the availability of these data, which were used under license for the current study and are not publicly available. Data may be made available by the authors upon reasonable request, with permission of Yogyakarta District Health Office.

## Acknowledgements

We gratefully acknowledge the contributions of the staff in the Disease Control Division at Yogyakarta City Health Office for providing surveillance data; staff at the Yogyakarta puskesmas clinics for providing rapid diagnostic test result data; community members for their support and participation, particularly their contribution to mosquito releases and collections; provincial, district and kelurahan leaders for their support for this project; and all members of the World Mosquito Program (WMP) Yogyakarta and Global teams who contributed to the planning and implementation of the quasi experimental study.

## List of abbreviations

AWED: ‘Applying *Wolbachia* to eliminate dengue’ (randomised controlled trial currently ongoing in Yogyakarta)
BG trap: Biogents Sentinel Trap
CI: confidence intervals
CRCT: cluster randomised controlled trial
DENV: dengue virus
DF: dengue fever
DHF: dengue haemorrhagic fever
DHO: District Health Office
ICD-10: International Classification of Diseases, 10^th^ revision
IRR: Incidence rate ratio
ITS: interrupted time series
MRC: Mosquito release container
NS1: non-structural protein
1 QA: quality assurance
RDT: rapid diagnostic test
RT-PCR: reverse transcription polymerase chain reaction
WMP: World Mosquito Program

---

**Figure S1.**
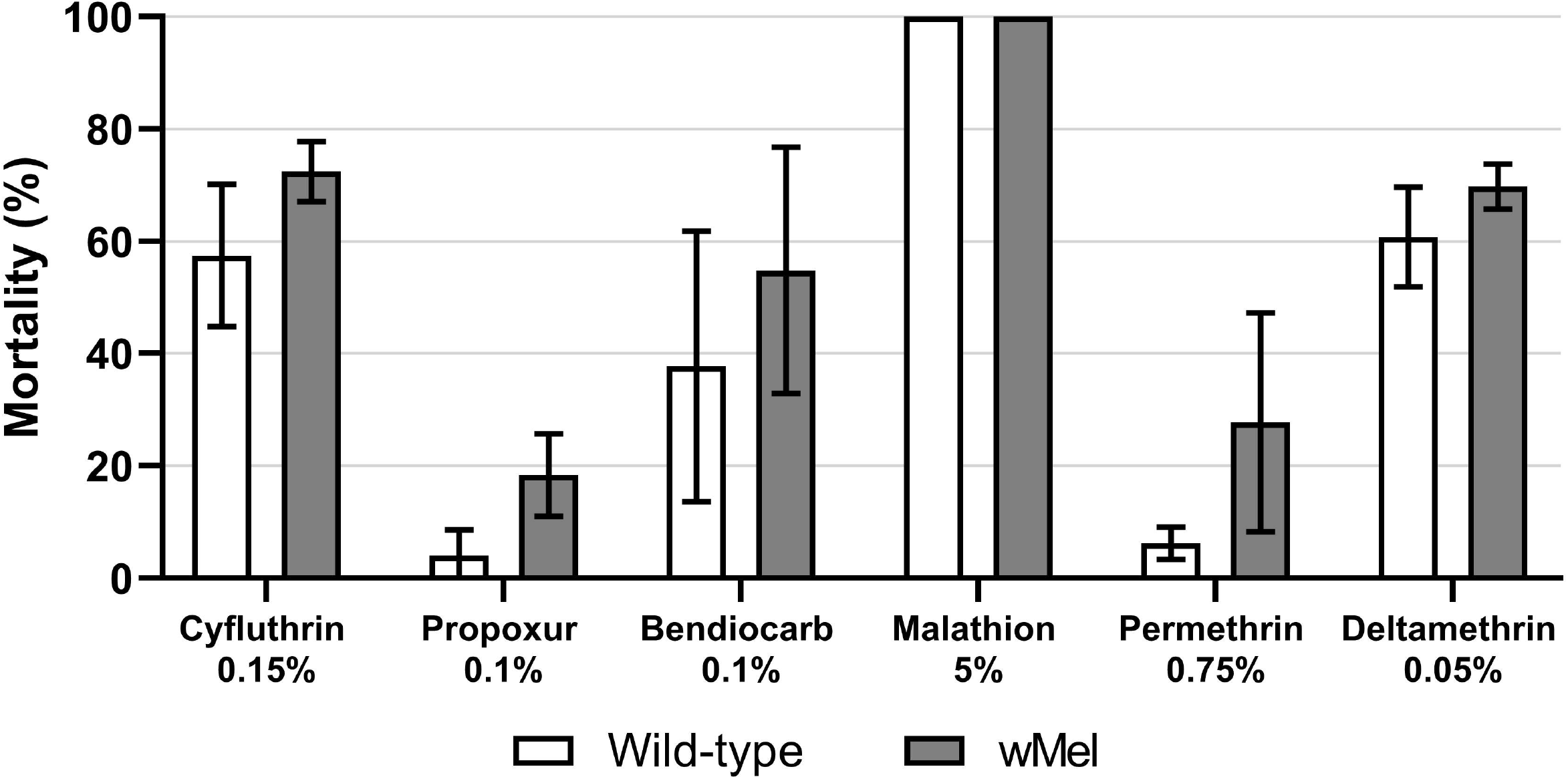
Insecticide susceptibility of the *w*Mel *Ae. aegypti* colony and wild-type *Ae. aegypti* collected from the quasi-experimental study intervention area. Bars show mean (st. dev.) % mortality across four replicate tests, each with 20 – 28 mosquitoes tested against each insecticide.

